# Explanations for higher-than-expected mortality from April 2021: a scoping review protocol

**DOI:** 10.1101/2023.06.20.23291333

**Authors:** Margaret Douglas, Gerry McCartney, David Walsh, Grant Donaghy, David Rae, Sarah Wild, Julie Ramsay

## Abstract

**Objective:** The objective of this scoping review is to identify the explanations that have been proposed for higher-than-expected mortality following the first pandemic year, and any evidence to support or refute these explanations.

**Introduction:** Mortality rates have remained high compared to previous years, beyond the peak waves of Covid-19 mortality. Several explanations have been suggested for this. Identifying potential hypotheses and empirical studies investigating these is the first step before any further analytical work to investigate these trends can be undertaken.

**Inclusion criteria:** The scoping review will include papers proposing or investigating hypotheses for raised all cause or cause specific mortality, or reduced life expectancy, from April 2021 onwards compared to pre-pandemic levels. It will include papers on mortality in the whole population or any specific demographic sub-populations, in high income countries only, but exclude studies of mortality or survival following a healthcare intervention.

**Methods:** A systematic search will be undertaken on Medline, Embase and Google Scholar for relevant articles published from 2021 onwards in English, with a similar search for grey literature on relevant government websites. Two reviewers will screen titles and abstracts, then full text articles with disagreements resolved by discussion or involvement of a third reviewer. Data extracted from selected articles will include the setting, population, hypothesis/es proposed, study type and findings if relevant. Included papers will be tabulated against the proposed hypotheses with any empirical evidence and hypotheses summarised narratively.

## Introduction

In the UK and other countries, there has been concern about excess mortality compared to previous years, continuing beyond the peak waves of mortality associated with the pandemic^1^. Some of these are attributable to Covid-19, which is still an important case of death, but there has also been a higher-than-expected number of non-covid deaths^2 3^.

Given the characteristics of people most vulnerable to dying directly from Covid-19, the pandemic is likely to have brought forward deaths in some people who would otherwise have died within a few years^4^, although the Years of Life Lost (YLL) directly from Covid-19 in Scotland early in the pandemic were still substantial^5^. This displacement effect would be expected to lead to mortality subsequently being *lower* than previous years, rather than the continuing higher rates that are observed.

There is debate about the appropriate baseline to define ‘excess’ mortality^6^. One common method has been to compare with the average mortality rate for the same week or month in the previous 5 years. It is not appropriate to include 2020 in the baseline as mortality was inflated by the first waves of Covid-19, but an alternative baseline can be used that excludes 2020^7^. As the population is ageing, changes in age structure in the past 5-6 years mean that comparisons of crude mortality counts or rates are likely to be misleading^1^. Age-standardised mortality rates provide a better comparison^8^ and, since 2021, have also shown an excess over pre-2020 rates. However, comparisons even of age standardised rates may be misleading when the population age structure has changed. Life expectancy is an alternative measure to show changes in mortality after accounting for changes in the population age structure.

There are several possible explanations for the continuing high mortality. These include: the longer term impacts of Covid-19 disease^9^; the effects of delayed or missed healthcare due to continuing health service pressures^1 10^, including ‘secondary prevention’ measures^11^; high rates of influenza and other respiratory infections that occurred following lifting of physical distancing measures; the wider impacts of the pandemic response operating through a range of economic and social pathways^10^, the impacts of wider political and societal pressures including the cost-of-living crisis^12^, the summer 2022 heatwave^13^, and continuation of the previously noted stalling or reversal of improvements in mortality that were of concern long before the pandemic^14^.

In the UK, death rates in more deprived populations were already increasing before the pandemic, and at the national level long-term improvements in mortality had stalled from around 2012^15 16^. This affected both men and women, almost all age groups and has been observed for many different causes of deaths. Similar trends have been seen, to different degrees, in other countries. A large body of research studying the reasons for these pre-pandemic trends concluded that these trends were associated with austerity policies introduced in the UK and other countries from the early 2010s ^14 17 18^. Many of the financial and social impacts of austerity have been exacerbated by the pandemic and the more recent cost of living crisis^19^.

A critical understanding of the reasons for continuing high mortality rates is important to inform policy responses. Before conducting analytical work to investigate the reasons, it is important to identify all the hypotheses put forward to explain the trends, and any analyses already completed to investigate these. A scoping review is an appropriate way to collate and map the literature on a topic. We carried out a preliminary search of MEDLINE, the Cochrane Database of Systematic Reviews and *JBI Evidence Synthesis* on 23^rd^ May 2023 and identified no current or underway systematic reviews or scoping reviews on the topic of mortality trends since 2021.

We therefore propose a scoping review that will aim to assess the extent of current literature theorising or explaining the continuing higher-than-expected mortality beyond the peak of Covid-19 deaths. The review will focus on high income countries and will seek to identify all the hypotheses that have been suggested for these trends, and to identify and critically appraise any analytical studies investigating these hypotheses.

### Review question

What explanations have been proposed for higher-than-expected mortality in high income countries persisting beyond the first year of the Covid-19 pandemic, and what empirical evidence is available either supporting or refuting these hypotheses?

### Eligibility criteria

#### Participants

Whole population or any specific sub-populations (e.g. children, maternal or people with long term conditions).

#### Concept

Proposed theories or evidence to explain worse-than-expected mortality or life expectancy (all-cause and cause-specific) from April 2021 onwards compared to pre-pandemic levels.

Exclusions: studies of mortality or survival following a healthcare intervention; studies comparing survival in people with and without a diagnosis; studies from countries that have not experienced worse than expected mortality from April 2021.

#### Context

High income countries.

From April 2021 onwards (As in most countries this is avoids the main mortality peaks caused directly by Covid-19).

#### Types of Sources

This scoping review will consider any papers and studies that propose and/or investigate reasons for higher-than-expected mortality trends. This will include text and opinion papers, including blogs, any analytical or observational study design, and any systematic reviews that meet the inclusion criteria and consider one or more explanation for mortality trends.

## Methods

The proposed scoping review will be conducted in accordance with the JBI methodology for scoping reviews^20^.

### Search strategy

The search strategy will aim to locate papers in both peer reviewed and grey literature.

An initial limited search of MEDLINE was undertaken to identify articles on the topic. The text words contained in the titles and abstracts of relevant articles, and the index terms used to describe the articles were used to develop a full search strategy for Medline, Embase and Scopus.

The search strategy, including all identified keywords and index terms, will be adapted for each included database and/or information source.

The reference list of all included sources of evidence will be screened for additional studies. The search will be restricted to papers published in English.

Papers published from 2021 onwards will be included as the focus is excess mortality trends continuing beyond the first pandemic year.

The databases to be searched include Medline, Embase and Google Scholar.

Sources of grey literature to be searched will include:

- National Records of Scotland
- Office for National Statistics
- Glasgow Centre for Population Health
- Public Health Scotland
- Scottish Government
- Healthcare Improvement Scotland
- UKHSA / OHID / UK Government
- King’s Fund
- Joseph Rowntree Organisation
- WHO
- NICE
- NHS England
- Public Health Wales
- Public Health Agency Northern Ireland
- The Health Foundation

### Study/Source of Evidence selection

Following the search, all identified citations will be collated, uploaded into Covidence and duplicates removed. Following a pilot test, titles and abstracts will then be screened by two or more independent reviewers for assessment against the inclusion criteria for the review. Potentially relevant sources will be retrieved in full. The full text of selected citations will be assessed in detail against the inclusion criteria by two or more independent reviewers. Reasons for exclusion of sources of evidence at full text that do not meet the inclusion criteria will be recorded and reported in the scoping review.

Any disagreements that arise between the reviewers at each stage of the selection process will be resolved through discussion, or by mediation by an additional reviewer/s.

The results of the search and the study inclusion process will be reported in full in the final scoping review and presented in a Preferred Reporting Items for Systematic Reviews and Meta-analyses extension for scoping review (PRISMA-ScR) flow diagram ^21^.

### Data Extraction

Data will be extracted from papers and entered into a bespoke data extraction form developed by the reviewers. The data extracted will include:

- Citation
- Dates of data presented
- Setting
- Metrics (mortality, premature mortality, life expectancy)
- Definition of expected mortality
- Population(s) (whole population, specified age group etc)
- Causes – all cause/specific causes
- Hypothesis/es proposed for mortality trends
- Study type (if includes empirical research)
- Findings (if any empirical research)

The reviewers will pilot the data extraction form on up to ten papers to identify other relevant variables and amend as required.

Data will be extracted by two reviewers for each paper. Any disagreements that arise between the reviewers will be resolved through discussion, or by involving an additional reviewer/s.

A draft extraction form is provided (see Appendix 2). The draft data extraction form will be modified and revised as necessary during the process of extracting data from each included evidence source. Modifications will be detailed in the scoping review. If appropriate, authors of papers will be contacted to request missing or additional data, where required.

### Critical appraisal of individual sources of evidence

Papers that simply put forward one or more hypotheses will not be critically appraised. Papers that contain any analysis testing one or more hypotheses will be critically appraised, using the relevant JBI checklist for the type of analysis (eg cross sectional analytical study, cohort studies or systematic reviews).

### Data Analysis and Presentation

Included papers will be tabulated against the hypotheses proposed as explanations for higher-than-expected mortality trends, with any empirical evidence provided in support these hypotheses. The hypotheses and any relevant evidence will be summarised narratively.

## Data Availability

This is a review protocol and no data have been produced

## Appendices

### Appendix I: Search strategy for Medline

1. “excess death*”.ti,ab.
2. “excess mortality”.ti,ab.
3. (“life expectancy” adj1 (lower or reduced)).ti,ab.
4. “increased death*”.ti,ab. 5
5. or/1-4
6. limit 5 to (english language and yr=“2021 -Current”)
7. “developing countries”.hw,kf,ti,ab.
8. (Africa* or Asia* or Caribbean or “West Indies” or “South America” or “Latin America” or “Central America”).hw,kf,ti,ab,cp. 828694
9. (Afghanistan or Algeria* or Argentina or Armenia* or Albania* or Angola* or Antigua or Azerbaijan or Bangladesh or Barbuda or Belarus or Benin or Belize or Bhutan or Bolivia* or Botswana or Bosnia* or Brazil* or “burkina Faso” or Burundi or “Cabo Verde” or Cambodia* or Cameroon or Congo or China or Chad or Colombia* or Comoros or “Cote d’Ivoire” or “Costa Rica” or “Central African Republic” or Cuba* or “Democratic Republic of the Congo” or Djibouti or “Dominican Republic” or Dominica* or Ecuador or “El Salvador” or “Equatorial Guinea” or Eritrea* or Eswatini or Ethiopia* or Egypt* or Fiji or Gabon or Gambia* or Georgia or Guinea or Ghana or Grenada or Guatemala or “Guinea-Bissau” or Guyana or Haiti or Herzegovina or Honduras or India* or Indonesia* or Iran or Iraq* or Jamaica* or Jordan or Kazakhstan or Kenya or Korea* or Kosovo or Kyrgyzstan or Kiribati or “Lao People’s Democratic Republic” or Lebanon or Lesotho or Liberia or Libya* or Macedonia* or Madagascar or Malaysia* or Malawi or Maldives or Mali or Mauritius or “Marshall Islands” or Mauritania or Mexic* or Micronesia or Moldova or Mongolia* or Montenegro or Montserrat or Morocco or Mozambique or Myanmar or Namibia or Nauru or Nepal* or Nigeria* or Nicaragua* or Niue or Niger or Pakistan or “Papua New Guinea” or Palau or Panama or Paraguay or Peru or Philippines or Rwanda* or “Sao Tome and Principe” or Senegal or “Saint Helena” or “Saint Lucia” or “Saint Vincent and the Grenadine” or Samoa or Serbia* or “South Africa*” or “Sri Lanka” or Suriname or “Sierra Leone” or “Solomon Islands” or Somalia* or Sudan* or Syria* or Tanzania* or Tajikistan or Tokelau or “Timor-Leste” or Thailand or Tonga or Turkey or Turkmenistan or Tunisia* or Togo or Tuvalu or “United Arab Emirates” or Ukrain* or Uganda* or Uzbekistan or Vanuatu or Venezuela* or Vietnam* or “West bank and Gaza Strip” or “Wallis and Futuna” or Yemen or Zambia* or Zimbabwe).hw,kf,ti,ab,cp.
10. ((developing or “less* developed” or “under developed” or underdeveloped or “middle income” or “low* income” or underserved or “under served” or deprived or poor*) adj3 (countr* or nation? or population? or world)).ti,ab.
11. ((developing or “less* developed” or “under developed” or underdeveloped or “middle income” or “low* income”) adj3 (economy or economies)).ti,ab.
12. (low* adj3 (gdp or gnp or “gross domestic” or “gross national”)).ti,ab.
13. (low* adj3 middle adj3 countr*).ti,ab.
14. (lmic or lmics or “third world” or “lami countr*”).ti,ab.
15. “transitional countr*”.ti,ab.
16. ((“high burden” or “high-burden” or countdown) adj3 countr*).ti,ab.
17. “global south”.ti,ab,kf,hw. 18 or/7-17
18. 19 6 not 18

### Appendix II: Draft data extraction instrument

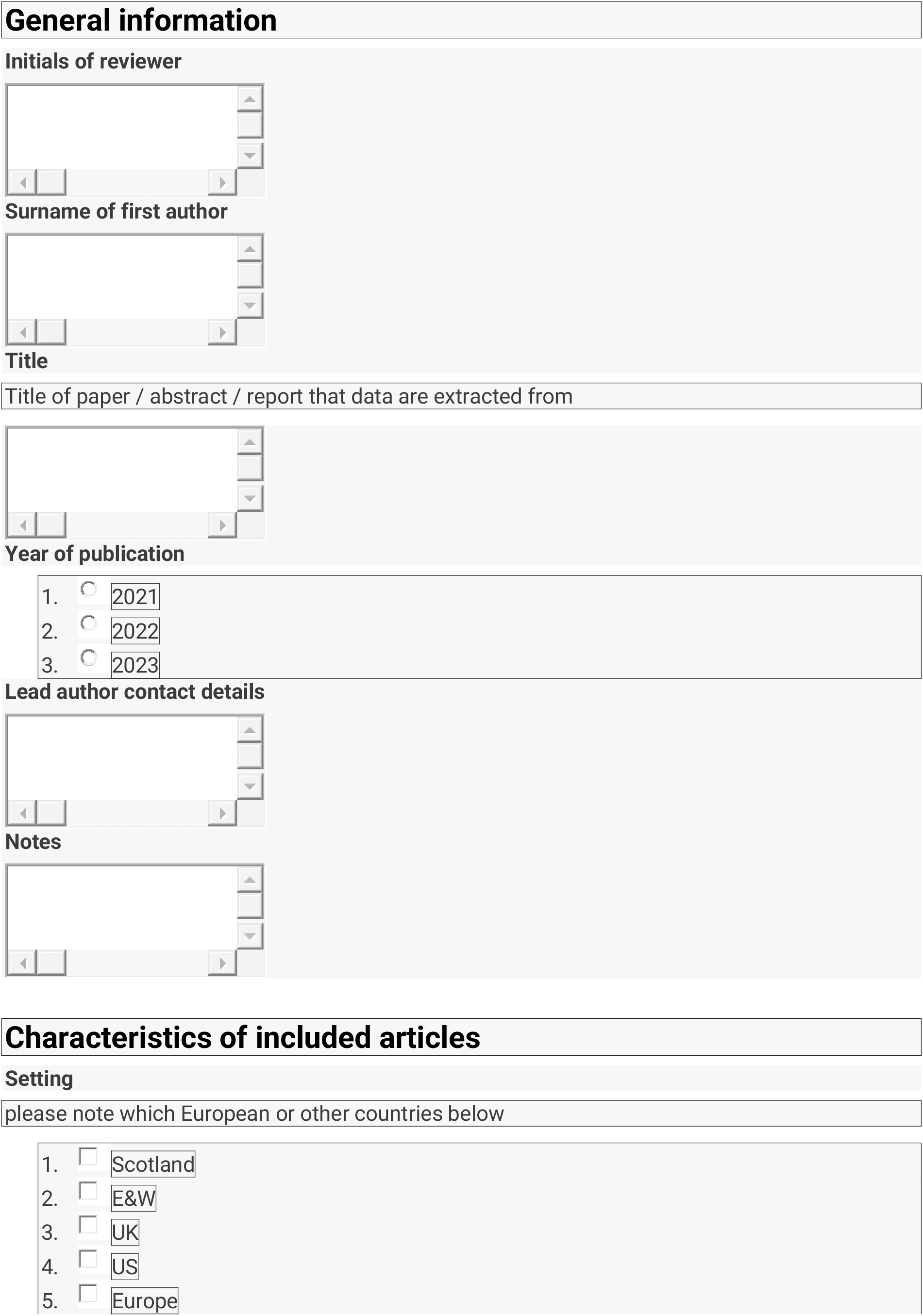

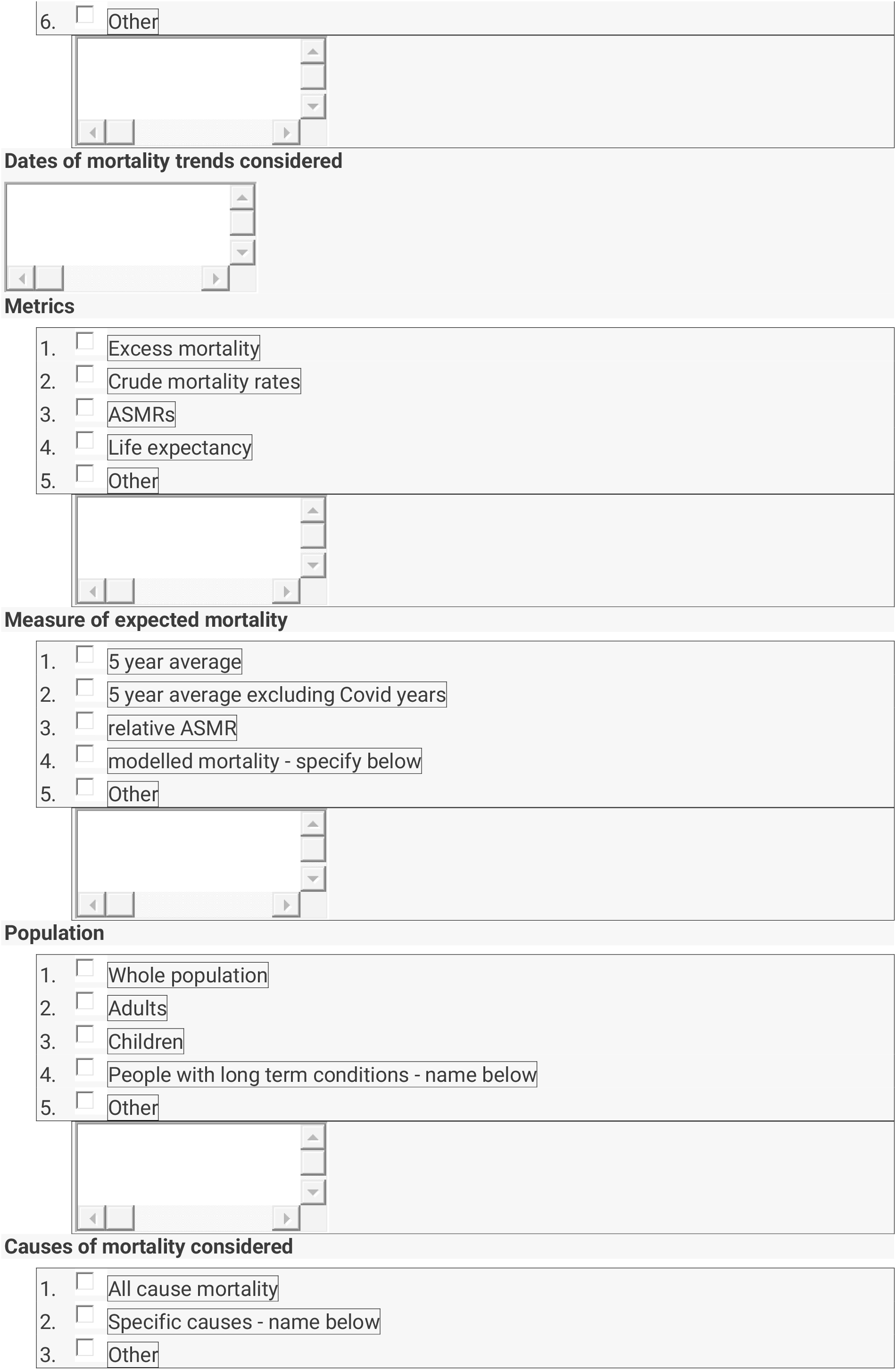

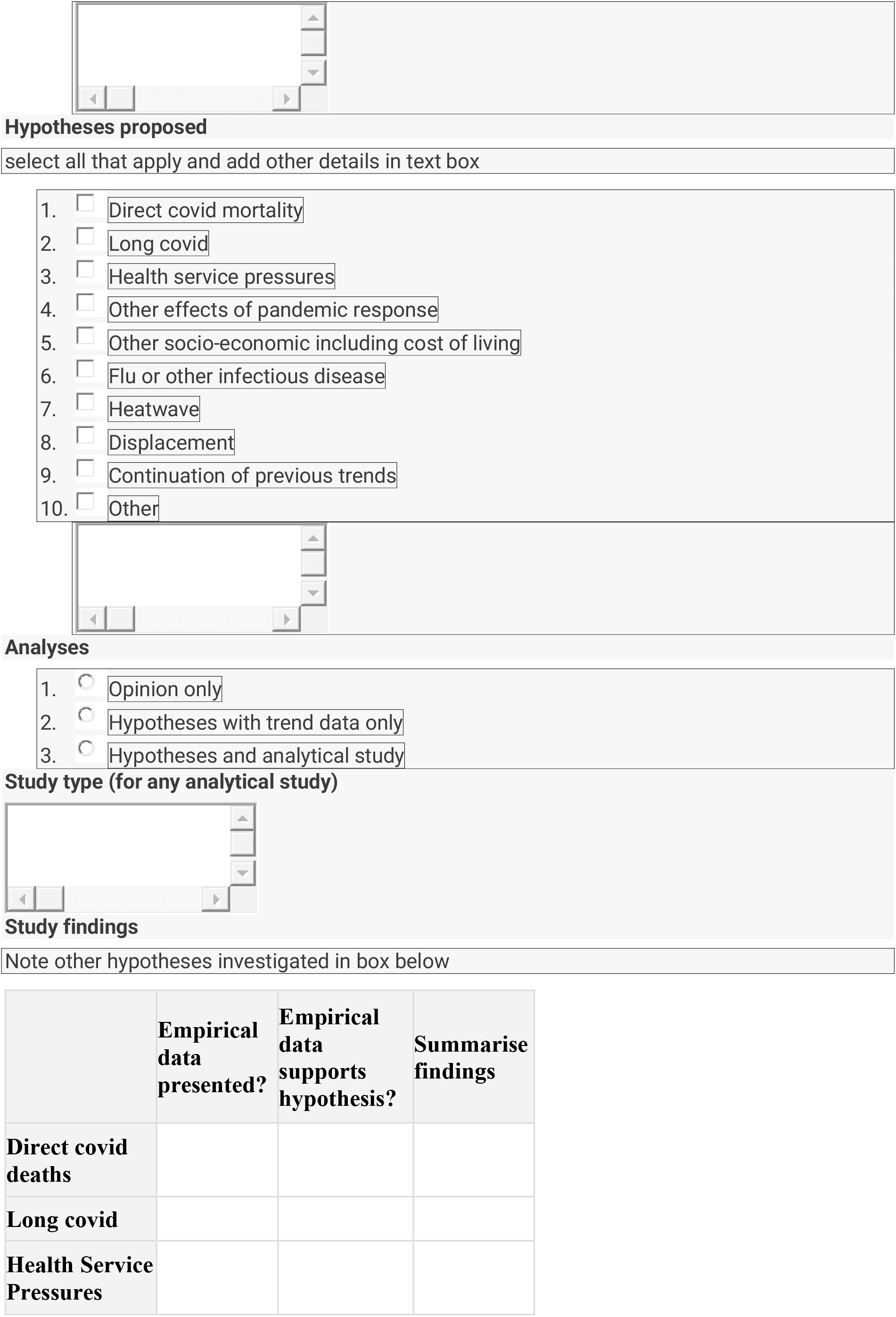

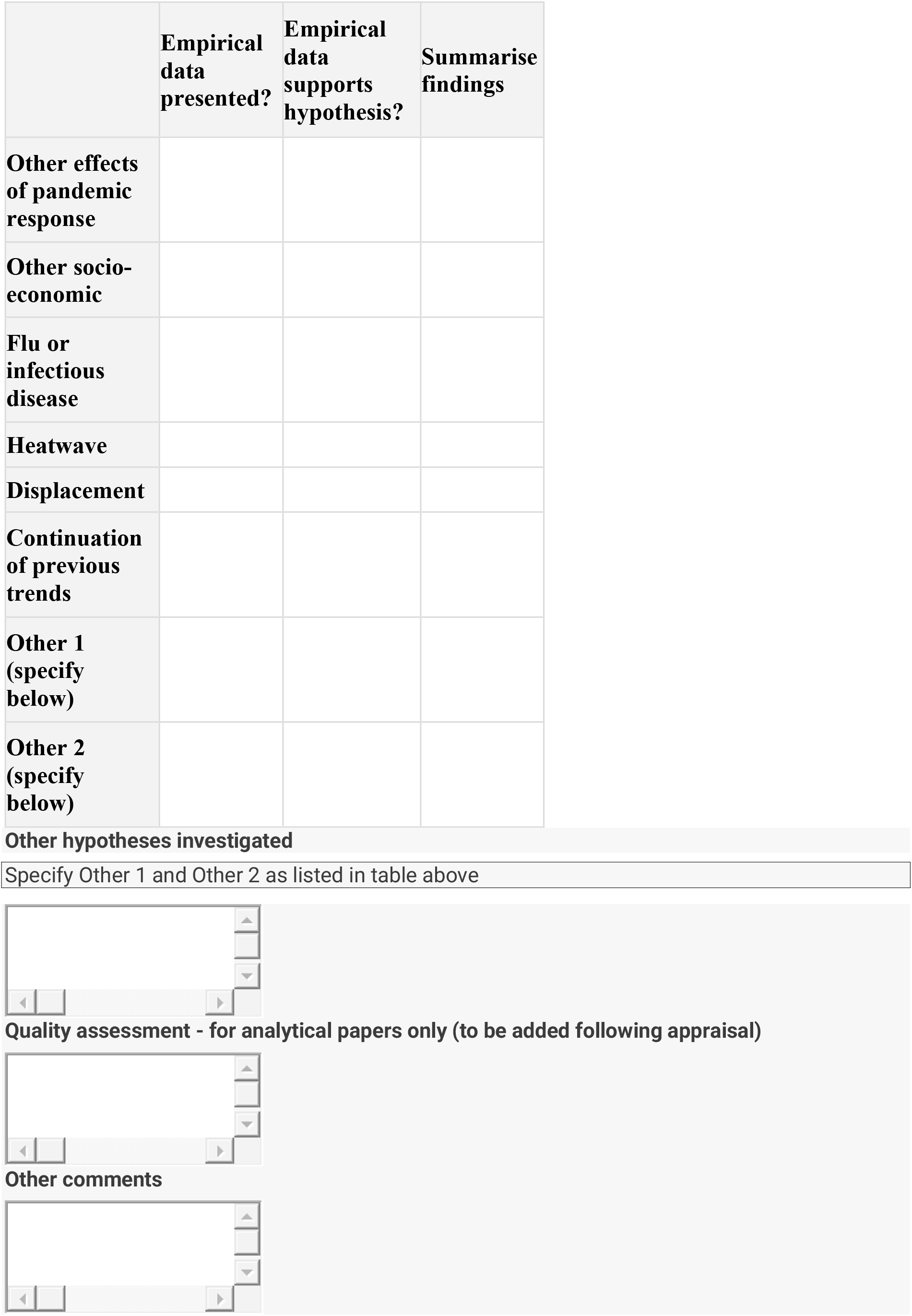

